# TrialCalibre1: An Agentic System for Trial Emulation, Benchmarking, and Calibration

**DOI:** 10.64898/2026.10.13.26365421

**Authors:** Amir Habibdoust, Xing Song

## Abstract

TrialCalibre1 is an agentic system that connects clinical evidence, study design, and executable analyses. It supports reference-trial replication, direct target-trial emulation, and BenchExCal, which combines benchmarking, extension, and calibration sensitivity analysis. We evaluated an ASCET-based benchmark and an extension comparing clopidogrel with aspirin among adults aged 81–90 years using synthetic PCORnet-style claims. Two reviewers assessed benchmark extraction, protocol decisions, and implementation. After discussion, 53/62 and 50/62 assessable SQL checks were rated met; both reviewers rated 12/13 assessable R checks met. Protocol and proxy disagreements remained. A separate ten-question routing development set yielded 10/10 route matches and 7/7 applicable reference-trial matches. The agent benchmark and extension included 6,069 and 1,602 patients. The extension risk ratio was 0.848 (95% CI 0.569–1.184), with a calibrated sensitivity estimate of 1.274 (0.439–3.700). The human implementation provided descriptive comparisons, not same-data validation. Dose proxies, unpopulated covariates, residual imbalance, implementation deviations, and ignored cross-cohort covariance limited interpretation. This case demonstrates a connected workflow whose protocols, code, and results can be reviewed.

## 1 Introduction and related work

Target-trial emulation (TTE) makes the intended randomized experiment explicit before translating it into observational data. Investigators specify eligibility, treatment strategies, assignment, followup, outcomes, causal contrast, and analysis, then determine which elements the available records can reliably capture (Hernán and Robins, 2016). Eligibility, assignment, and follow-up must be aligned in time, with attention to confounding and the timing, severity, and treatment details captured by the records (Dahabreh et al., 2025).

These difficulties also affect human-designed studies. Among 237 published TTE studies, Ren et al. (2026) found that 43.5% omitted at least one of the seven core components from their reports and 15.2% used postbaseline information inappropriately to determine eligibility. A review of 24 software tools identified uneven support for early design and interoperability across the workflow (van Hal et al., 2025). Agentic systems may help connect these tasks, but their evaluation must distinguish plausible prose, faithful implementation, numerical correctness, and credible causal inference.

Related systems offer complementary approaches to trial emulation. Trial Pathfinder evaluates how eligibility restrictions influence oncology populations and estimates (Liu et al., 2021). TRI-ALSCOPE connects clinical-data processing, trial-specific cohorts, and weighted survival analysis (González et al., 2025). EmulatRx coordinates specialized agents, terminology grounding, criterion-level SQL, execution feedback, and expert review (Li et al., 2026). THESEUS translates reviewed study specifications into executable analytical settings through deterministic rules (Kim et al., 2026). Automated workflows still require transparent assumptions and causal expertise (Guan et al., 2026).

TrialCalibre1 extends the TrialCalibre framework for observational benchmarking and calibration (Habibdoust and Song, 2025). It links the research question to approved operational definitions, terminology, cohort construction, and analysis. Its three pathways distinguish replication of a suitable trial, direct emulation without a suitable reference, and BenchExCal. The latter benchmarks against a randomized trial, expands to a related question, and uses benchmark divergence to inform a calibration sensitivity analysis (Wang et al., 2025). Extension validity depends on justified assumptions about the transferability of divergence, even when the benchmark agrees with the trial.

We examine one retrospective synthetic-data case involving an older population outside ASCET’s eligibility, together with a ten-question router evaluation. A human-designed benchmark, extension, and calibration provide another implementation of the question. The contribution is a reviewable link between study decisions and executable outputs, not a new causal estimator, a clinical treatment recommendation, or evidence of superiority over existing systems. For clinical research, the key question is whether the implemented comparison matches the intended population, treatment decision, and time horizon, not simply whether the generated text reads well.

## 2 System architecture

TrialCalibre1 combines specialized language-model components with deterministic workflow control and local execution (Figure 1). A study-design router proposes one of the three pathways and any reference trial for approval. The Protocol Builder receives the question, reference evidence, and an initial assessment of database metadata, then specifies eligibility, treatment strategies, time zero, outcomes, follow-up, estimand, covariates, and analysis requirements. Reference definitions remain distinct from observational adaptations. Missing measurements lead to explicit proxy proposals or omission decisions, with unresolved scientific choices reviewed before implementation.

**Figure 1.**
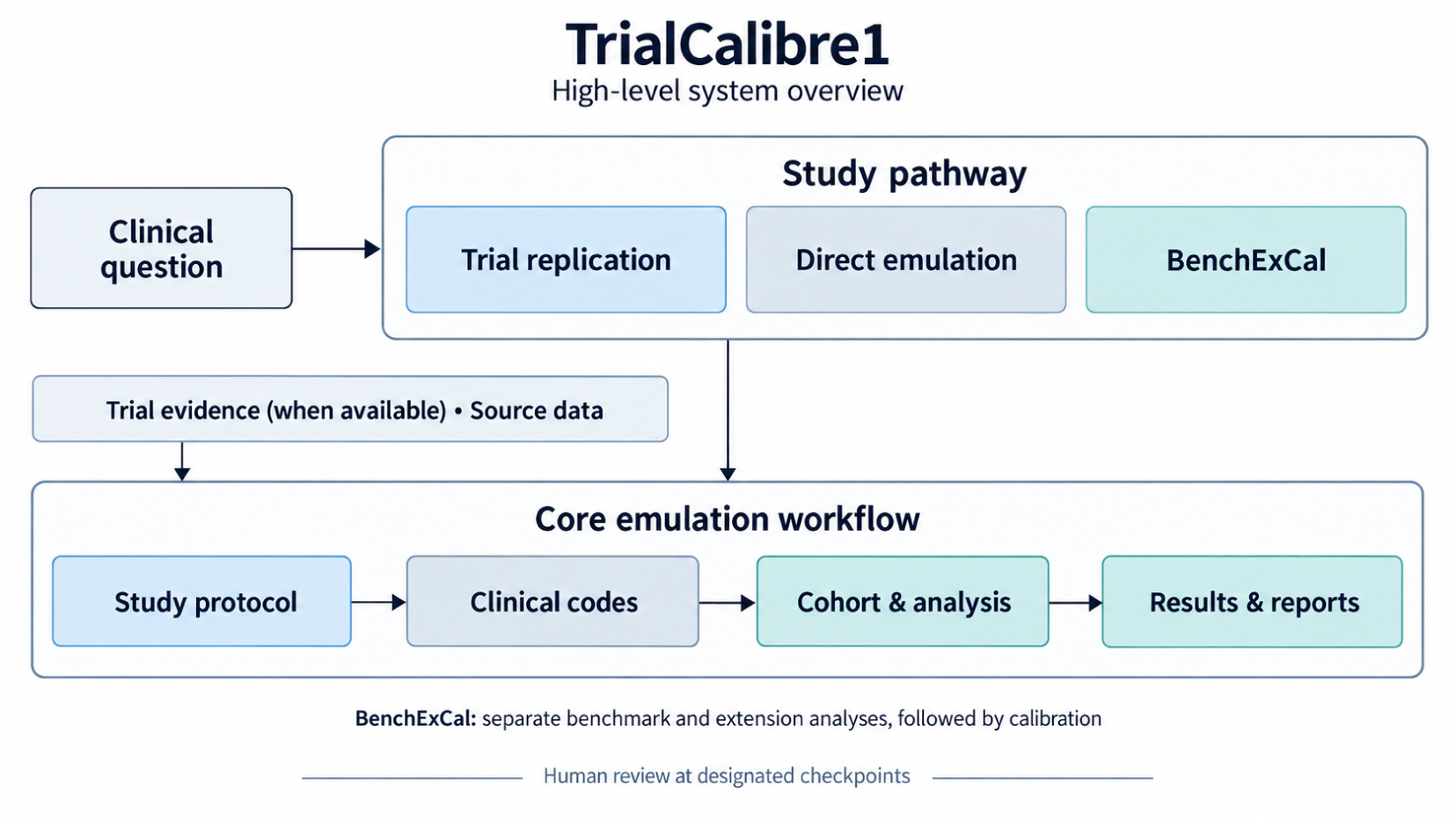
TrialCalibre1 study pathways and shared workflow. Replication, BenchExCal, and direct emulation share protocol development, coding, cohort construction, and reporting. BenchExCal retains separate benchmark and extension analyses. Human review occurs at designated checkpoints. The schematic was generated using OpenAI’s image-generation tool.

The local secure-execution component identifies table and column names, data types, keys, relationships, and coding systems. The architecture is designed to keep patient-level records within this environment; downstream LLM agents receive metadata, execution diagnostics, and restricted aggregate summaries rather than individual patient records. This information helps assess protocol feasibility, select proxies, and write SQL. Measurement adequacy requires assessment beyond the presence of a field. TrialCode Agent, a separately developed and evaluated terminology component, supplies curated definitions or new candidates with documented sources (Habibdoust et al., 2026). Database coverage requires checking whether valid codes match observed records.

The cohort-construction component translates the approved emulation protocol into SQL and produces the analysis dataset.

For supported analyses, model-proposed settings configure a fixed statistical engine that produces R. Descriptive adherence and switching reports follow the main estimates, with baseline assignment unchanged. BenchExCal preserves the benchmark results and produces distinct extension and calibration reports. The evaluated implementation uses SQLite and a two-treatment statistical engine. Saved protocols, decisions, code, and outputs support review.

## 3 Case study and evaluation methods

### 3.1 Clinical question and data

The submitted question asked: among adults aged 81–90 years with documented coronary heart disease already receiving aspirin, what is the effect of switching to clopidogrel 75 mg/day versus continuing aspirin 162 mg/day on the two-year composite risk of unstable angina, myocardial infarction, stroke, or death? Requested outputs were arm-specific risks, risk difference (RD), risk ratio (RR), and number needed to treat or harm (NNT/NNH), with confidence intervals.

The approved BenchExCal pathway used ASCET as its benchmark. ASCET randomized 1,001 aspirin-treated patients to clopidogrel 75 mg/day or aspirin 160 mg/day; its eligibility extended to age 80 and its composite specified nonhemorrhagic stroke (Pettersen et al., 2012). The requested older population, aspirin dose, and broader stroke wording were therefore distinct from the reference. The synthetic database was generated and enriched using custom Python scripts and stored in SQLite following PCORnet Common Data Model-style table conventions (Waitman et al., 2014, 2022). It contained demographics, enrollment, dispensing, diagnoses, procedures, encounters, and deaths, but no confirmed laboratory or vital-sign measurements. No causal ground truth was established.

Both workflows used prior aspirin exposure to distinguish the question from an ordinary new-user comparison. Candidate treatment dates allowed eligibility to vary over time; each final cohort retained one earliest eligible entry per patient. The analysis used intention-to-treat-like baseline assignment and follow-up capped at 730 days. Because a suitable NDC code for aspirin 160 mg was unavailable for this test, the reviewer directed the agent to use an 81-mg product proxy; the human emulation used the same proxy. The human analysis explicitly assumed one 81-mg tablet daily. These analyses address the implemented aspirin definitions, with daily-dose equivalence unresolved.

### 3.2 Evaluation scope and human comparison

We organize this retrospective feasibility and artifact-review evaluation around reviewer-assessed fidelity of saved implementations to approved study specifications. Protocol/proxy judgments, targeted terminology findings, and router agreement are reported separately. Cohort counts and treatment estimates demonstrate workflow outputs; they do not establish patient-level cohort accuracy, independent numerical correctness, causal-effect recovery, or general reliability. The ASCET-based benchmark, older-age extension, and calibration form one case; the ten-question router assessment is a separate development-set evaluation.

The extension adapted the benchmark definitions to the older population. GLM 5.2 was used for benchmark source extraction, protocol development, and the terminology component. Gemini 3.7 Flash with low reasoning was used for SQL generation, statistical configuration, extension protocol adaptation, randomized-reference extraction for calibration, and the ten-question router evaluation. Both models were accessed through OpenRouter. Evaluation focused on the final protocols, SQL, R, cohorts, and estimates; end-to-end autonomous performance, runtime, and cost remain targets for future evaluation.

One investigator implemented a human-designed benchmark and older-age extension. Its age extension changed the inclusive eligibility range from 18–80 to 81–90, rebuilt affected downstream tables, and refitted the same analysis specification. Its eligibility, code coverage, outcomes, observation rules, and weighting differed from the agent’s. Whether the source databases contain identical underlying records remains unconfirmed. This comparison is not a patient-level gold standard, a blinded experiment, or independent numerical verification on the agent’s frozen analysis dataset.

### 3.3 Protocol and implementation review

Two human reviewers assessed benchmark trial extraction, protocol, proxies, covariates, SQL, and R. Review workbooks included trial/emulation definitions and a SQL checklist with prefilled requirement/evidence columns; one reviewer cleared those columns before scoring. Reviewer-directed changes followed discussion; retained R1/R2 scores are reported separately as post-discussion judgments, with consensus and independent pre-discussion inter-rater reliability left undetermined (Kottner et al., 2011).

Review domains follow target-trial specification and reporting principles (Hernán and Robins, 2016; Cashin et al., 2025). Extraction ratings were correct/incorrect; protocol and proxy rubrics distinguished appropriate or acceptable definitions from revisions and missing or unacceptable elements. Covariates were rated relevant/not relevant. Proxy judgments also covered omissions and adaptations, so their acceptable proportion describes reviewer acceptance rather than diagnostic validity. N/A, not-needed, no-proxy, and unreviewed entries remain distinct.

Terminology generation relied on TrialCode Agent, evaluated in prior work (Habibdoust et al., 2026). We did not conduct a comprehensive independent code-by-code accuracy assessment. SQL review assessed consistent use of the approved sets, rather than independently validating their clinical accuracy. A targeted inspection of constant covariates examined the final analysis data and saved code mappings.

SQL review compared executed scripts and saved diagnostics with the approved protocol, amendments, codes, and execution plan. It did not require reviewers to rewrite SQL. Checks addressed individual criteria, eligibility before index selection, assignment, time zero, outcomes, censoring, covariates, and relational integrity. Error categories included incorrect logical connections, missing concepts, integrity violations, and schema errors (Li et al., 2026). Execution alone cannot establish semantic correctness (Zhong et al., 2020); we did not apply their test-suite method here.

SQL and the 14-item R review used met/not met/unclear/N/A ratings. Fidelity was met/(met + not met), with unclear, N/A, and unreviewed counts reported separately. These study-specific checks are not independent defects or patient-level accuracy measures.

The R review covered inputs, treatment coding, propensity scores, weighting, balance diagnostics, risks, contrasts, uncertainty, supplementary hazard ratios, and additional analyses. It was code/specification review supported by saved outputs, not independent re-estimation. Such same-data verification remains a separate task, consistent with validation principles based on independent calculations or trusted implementations (Lee et al., 2026). One human reviewer confirmed 15 agent BenchExCal checks: six extension protocol/SQL, four randomized-reference, and five calibration checks. This assessment used saved evidence without rerunning the cohort or statistical analysis. Two-reviewer assessment is confined to the benchmark.

### 3.4 Statistical analysis and calibration

The agent’s recorded plan labeled the Cox model primary; this paper reports the requested risk-based contrasts as primary. Each age cohort used propensity-score weighting, weighted Kaplan–Meier risks, and 1,000 patient bootstrap samples with propensity-score re-estimation. The agent used clipped propensity scores and stabilized, percentile-capped weights; the human workflow used a main-effects model with 18 covariates and unstabilized, untruncated average-treatment-effect weights. Bootstrap quantile conventions differed. Supplementary Cox estimates used robust variance-based intervals. When an RD interval crosses zero, its reciprocal NNT/NNH confidence set is unbounded across benefit and harm.

Calibration used bootstrap standard errors of the benchmark and extension log risk ratios. The common randomized reference was derived from ASCET’s first-event counts, 52/499 clopidogrel and 54/502 aspirin participants, assuming complete two-year ascertainment (Pettersen et al., 2012). It is an unadjusted count-based RR, not a published hazard ratio or censoring-adjusted estimate. Following BenchExCal, standardized benchmark-minus-reference divergence was subtracted on the log-relative-effect scale (Wang et al., 2025). This adapts benchexcal’s hazard-ratio implementation to RRs (Xue, 2026); Table S8 reports the calculation inputs.

Primary calibration used working-normal uncertainty and omitted benchmark–extension covariance despite 102 shared agent patients and 84 shared human patients. Approval authorized an exploratory approximation, not an independence claim. A human sensitivity analysis used 1,000 joint patient-cluster bootstrap samples with propensity refitting to estimate covariance, holding the estimated scaling factor fixed. These retrospective calculations were not prospectively registered or outcome-blinded and do not establish that divergence transfers between populations.

### 3.5 Router evaluation

A separate ten-question test contained four replication, three BenchExCal, and three direct-emulation reference labels. An answer key based on the reference sources was set before the final test. Replication included narrower questions within a trial’s scope; BenchExCal required a suitable benchmark plus a new population, treatment strategy, formulation, or dose. Direct emulation applied when the focused search found no suitable trial. The router used Gemini 3.7 Flash with low reasoning and received only the clinical questions; expected labels and reference answers were withheld. Recommendations were scored before human confirmation. Route agreement used all ten questions, while trial agreement used the seven applicable registry identifiers. All ten questions were evaluated together as a development set, not a held-out benchmark (Table S6). Direct-emulation labels remain conditional on the search.

## 4 Results

### 4.1 Cohorts and uncalibrated estimates

The approved pathway connected a 6,069-patient benchmark to a 1,602-patient older-age extension (Table 1). The benchmark produced 17 main SQL files: eight cohort-processing roles and nine individual eligibility tables, covering four inclusion and five exclusion criteria. It retained 595,308 candidate rows, 33,549 inclusion-positive candidate rows, and one final record per patient. The human cohorts contained 5,299 and 1,486 patients. Candidate rows are potential entries, not distinct patients. Patient-level overlap between the agent and human workflows was not calculated, so cohort-count differences are not false-positive or false-negative agent records.

**Table 1.** Agent and human cohort sizes and primary estimates.

| Measure | Agent 18–80 | Human 18–80 | Agent 81–90 | Human 81–90 |
| --- | --- | --- | --- | --- |
| Patients, n | 6,069 | 5,299 | 1,602 | 1,486 |
| Aspirin / clopidogrel, n | 3,827 / 2,242 | 3,278 / 2,021 | 1,234 / 368 | 1,007 / 479 |
| First composite events, n | 852 | 523 | 212 | 153 |
| Aspirin risk, % | 15.43 | 11.66 | 14.33 | 11.33 |
| Clopidogrel risk, % | 12.78 | 8.96 | 12.15 | 11.21 |
| RD, percentage points | –2.65 (–4.52 to | –2.70 (–4.48 to | –2.18 (–6.67 to | –0.12 (–3.88 to |
| (95% CI) | –0.80) | –0.84) | 2.35) | 3.80) |
| RR (95% CI) | 0.828 | 0.768 | 0.848 | 0.990 |
|  | (0.718–0.946) | (0.637–0.923) | (0.569–1.184) | (0.696–1.376) |
| Calibrated RR (95% sensitivity interval) | N/A | N/A | 1.274 | 1.554 |
|  |  |  | (0.439–3.700) | (0.659–3.662) |

The agent and human benchmark RDs were −2.65 and −2.70 percentage points, respectively. These are descriptive comparisons across different cohorts and specifications, not evidence of implementation accuracy. Benchmark NNTs were 37.8 (95% CI 22.1–124.9) and 37.0 (22.3–119.6). Among varying agent benchmark covariates, the maximum weighted absolute standardized mean difference (SMD) was 0.065; effective sample size was 5,829.2. However, nine of 18 benchmark covariates and ten of 18 extension covariates were zero for every patient in both arms (Table S2). The calculation reported zero SMD for constant columns, which provides no evidence of successful adjustment for those covariates. Both age cohorts completed 1,000 bootstraps in each workflow. Agent estimates were reported as exploratory results.

In the older cohort, the agent NNT point estimate was 45.9, with an unbounded benefit/harm confidence set because the RD interval crossed zero. Both uncalibrated extension RR intervals included one. The agent retained weighted imbalance in the male-sex indicator (SMD 0.220) and encounter count (0.421). Four human extension covariates exceeded 0.10, with maximum 0.387. Human censoring before day 730 was 23.54% in the aspirin arm and 2.09% in the clopidogrel arm. These findings limit interpretation of both analyses.

### 4.2 Protocol and implementation findings

Benchmark extraction received mostly favorable ratings, while protocol and proxy judgments were less favorable and differed between reviewers (Table 2). Both reviewers considered all 18 recorded covariates relevant. Remaining disagreements involved one extraction item, six proxy/adaptation items, eight emulation items, and three SQL checks. These include substantive judgments and differences involving non-scored categories, with possible overlap between the underlying issues. R2 left overall protocol consistency unreviewed.

**Table 2.** Human review of agent outputs.

**A. Benchmark ratings retained by two reviewers after discussion**
| Domain and highest category | R1, n/N (%) | R2, n/N (%) |
| --- | --- | --- |
| Extraction: Correct | 17/17 (100.0) | 16/17 (94.1) |
| Proxy/adaptation: Acceptable | 12/13 (92.3) | 8/14 (57.1) |
| Emulation protocol: Appropriate | 14/18 (77.8) | 10/17 (58.8) |
| Covariate relevance: Relevant | 18/18 (100.0) | 18/18 (100.0) |
| SQL fidelity: Met | 53/62 (85.5) | 50/62 (80.6) |
| R fidelity: Met | 12/13 (92.3) | 12/13 (92.3) |

**B. Agent BenchExCal assessment confirmed by one human reviewer**
| Domain | Met | Not met | Unclear |
| --- | --- | --- | --- |
| Extension protocol/SQL | 3 | 2 | 1 |
| Randomized-reference alignment | 3 | 0 | 1 |
| Calibration implementation | 5 | 0 | 0 |

Panel A reports reviewer-specific post-discussion ratings, not consensus or independent agreement. Protocol denominators include substantive scored categories; SQL/R denominators are Met + Not met. Each reviewer additionally recorded four Unclear and two N/A SQL judgments and one Unclear R judgment. Panel B totals 11/15 Met, 2/15 Not met, and 2/15 Unclear, covering all 15 checks. Additional benchmark rating categories appear in Table S10.

The three SQL disagreements concerned life-limiting malignancy, compliance-limiting conditions, and timing of documented coronary heart disease. Both reviewers rated follow-up-end status and outcome ascertainment within follow-up boundaries as unclear. The identical R ratings included a missing proportional-hazards assessment for the supplementary Cox estimate and uncertainty about additional planned analyses.

In the extension assessment, the two not-met items concerned accepting CHD diagnosis or angiography instead of the required combination, and omitting outcome-setting/diagnosis-position restrictions and an administrative-data-end cap. Encounter scope and stroke-subtype alignment remained unclear. Patient-level effects of these deviations remain unquantified. All five calibration checks met the selected computational specification, including the approved covariance approximation.

### 4.3 Calibration and routing results

The common count-derived ASCET reference was RR 0.969 (95% CI 0.676–1.389). Calibration shifted the agent extension RR from 0.848 to 1.274 and the human RR from 0.990 to 1.554, with both sensitivity intervals spanning one (Table 1). The human covariance sensitivity gave RR 1.554 (0.656–3.681); no corresponding saved agent result was available. Calibrated treatment-effect estimates were limited to relative-risk contrasts and their sensitivity intervals. The 102 and 84 shared patients occurred across age cohorts, not simultaneously across treatment arms.

The router matched all ten predefined routes: 4/4 replication, 3/3 BenchExCal, and 3/3 direct emulation. Accepted reference trials were selected in all seven applicable cases. These recommendations show that routing worked on this development set. They do not establish held-out performance, general search recall, or successful downstream execution.

The appendix contains agent and human protocol mappings; weighting and integrity diagnostics (Table S1); covariate balance and definitions (Tables S2–S3); calendar entry and follow-up (Table S4); medication-use summaries (Table S5); router questions and responses (Table S6); additional estimates and calibration analyses (Tables S7–S8); cohort-construction counts (Table S9); and additional review-rating categories (Table S10).

## 5 Discussion

The case follows a connected workflow from an older-population question through benchmark emulation using ASCET as the reference trial, extension, and exploratory calibration. TrialCalibre1 saved protocols, codes, queries, and results for review throughout the workflow. These records let reviewers compare intended definitions with executed code and identify disagreements. The effects of this organization on reviewer workload, accuracy, and time remain to be evaluated. Related systems, including EmulatRx, offer complementary approaches to agentic trial emulation (Li et al., 2026).

Covariate construction limited confounding control. The nine all-zero benchmark covariates covered prior unstable angina, stroke, PCI, CABG, hyperlipidemia, heart failure, and statin, beta-blocker, and ACE inhibitor/ARB use; the extension’s chronic kidney disease flag was also zero for every patient. The prior-unstable-angina set contained only ICD-9-CM 411.1, while the ACE inhibitor/ARB set contained angiotensin II vasopressor products (Table S3). The ACE inhibitor/ARB set contains a therapeutic-class mapping error; the causes of the remaining all-zero flags are unresolved and may involve code coverage, implementation, or the synthetic data. Together with the observed residual imbalance, these findings preclude treating the reported estimates as evidence of adequate confounding control. The estimates illustrate workflow outputs, and the effect of correcting the definitions remains unquantified.

The human workflow is a non-equivalent descriptive comparator, not a reference standard. After inclusion, 6,424 agent and 6,535 human patients qualified at least once, with final cohorts of 6,069 and 5,299. Differences included CHD eligibility logic, an additional human 180-day clopidogrel washout, medication-code coverage, and contraindication definitions. Study-drug exclusions flagged at least one candidate date for 130 agent and 2,384 human patients; human proxies included a stent within 365 days and selected liver-disease diagnoses. These flags do not necessarily imply final exclusion, because a patient could qualify later, and cannot attribute the 770-patient cohort difference to a single rule. Each calibration uses its own benchmark divergence and uncertainty. Agreement or disagreement between these differently specified workflows therefore does not establish either implementation’s accuracy.

Calibration introduces additional assumptions rather than eliminating these concerns. The selected calculation transfers standardized divergence from a benchmark to an older population and adapts a hazard-ratio implementation to risk ratios. Its randomized input assumes complete ascertainment, and primary intervals omit dependence between age cohorts. The human covariance sensitivity addresses one uncertainty approximation, holding the estimated scaling factor fixed; estimating agent covariance and addressing residual confounding remain outstanding tasks. Transfer of benchmark divergence remains an assumption (Wang et al., 2025).

The evaluation is limited to one retrospective synthetic clinical example, with no established causal ground truth. It does not support prescribing conclusions, population-level performance estimates, or comparative efficiency claims. The ten-question routing set is small and development-informed; repeat-run stability remains to be evaluated. Execution evidence is limited to SQLite, this claims-based setting, and a two-treatment analysis engine; direct-emulation execution and other data models remain untested here.

The review also has limits. Two reviewers evaluated the benchmark, but discussion preceded the retained scores and disagreements remain. Assessing independent pre-discussion reliability requires ratings collected before discussion. Reviewers found some checklist sub-items unclear, including definition versus timing checks. This ambiguity may have contributed to rating differences, along with substantive disagreements. A single reviewer assessed the agent extension and calibration, and the human comparison was implemented by one investigator. Comprehensive assessment of patient-date boundaries requires testing beyond code review and saved diagnostics. Some exclusion criteria returned zero positive candidates, leaving their clinical correctness dependent on further assessment. Confirming identical source records across workflows and completing independent same-data numerical verification of the agent remain outstanding tasks. The unclear outcome/follow-up judgments and observed extension deviations remain relevant even where statistical-code ratings were favorable.

Further evaluation should lock application versions, data, approved specifications, and analysis settings before testing multiple studies and repeated runs. Evaluation of at least 10 additional studies is underway; their results are not included in this report. Held-out routing should be evaluated separately from downstream execution. Small hand-checked boundary cases can test temporal rules or criteria poorly represented in the main data. Independent calculations on the same frozen analysis dataset would test numerical correctness more directly than comparison with a differently designed cohort (Lee et al., 2026). BenchExCal studies should additionally assess dependence-aware uncertainty and whether benchmark divergence plausibly transfers to the extension. Reviewer-specific judgments should remain visible alongside any later adjudication.

## 6 Conclusion

TrialCalibre1 connects scientific specification, terminology, incremental cohort construction, supported analysis, and reporting across replication, direct emulation, and BenchExCal. The ASCET-based synthetic case completed a benchmark, older-age extension, and exploratory calibration, with a human-designed workflow and reviews revealing differences beyond those visible in final estimates alone. The findings support further evaluation of protocol-centered automation. Additional studies and independent same-data checks are needed to establish how consistently the workflow preserves scientific intent.

## Responsible use

TrialCalibre1 is intended for research, not individual treatment recommendations. This synthetic-data case does not justify prescribing, policy, or regulatory decisions. Measurement error, confounding, imbalance, and dependence can produce misleading evidence, particularly for poorly represented groups. Real-world use requires independent review of protocols and code, validation of calculations and calibration assumptions, subgroup assessment, and explicit reporting of unresolved limitations. Applications using real patient records require privacy, security, and data-governance controls, including preventing identifiable information from reaching unapproved external services.

## Data Availability

All data produced in the present study are available upon reasonable request to the authors

## Footnotes

40th Conference on Neural Information Processing Systems (NeurIPS 2026). Workshop: WMHS.

## TrialCalibre1 Supplementary Material

Target trial emulation and BenchExCal

Supporting protocols and results for the ASCET benchmark (ages 18–80) and older-age extension (81–90). Agent and human analyses are labeled separately.

### S1. Agent trial-to-emulation mapping

This unscored mapping records the agent’s intended protocol. Trial information reflects its extraction; gaps in the captured evidence do not establish omissions in the publication.

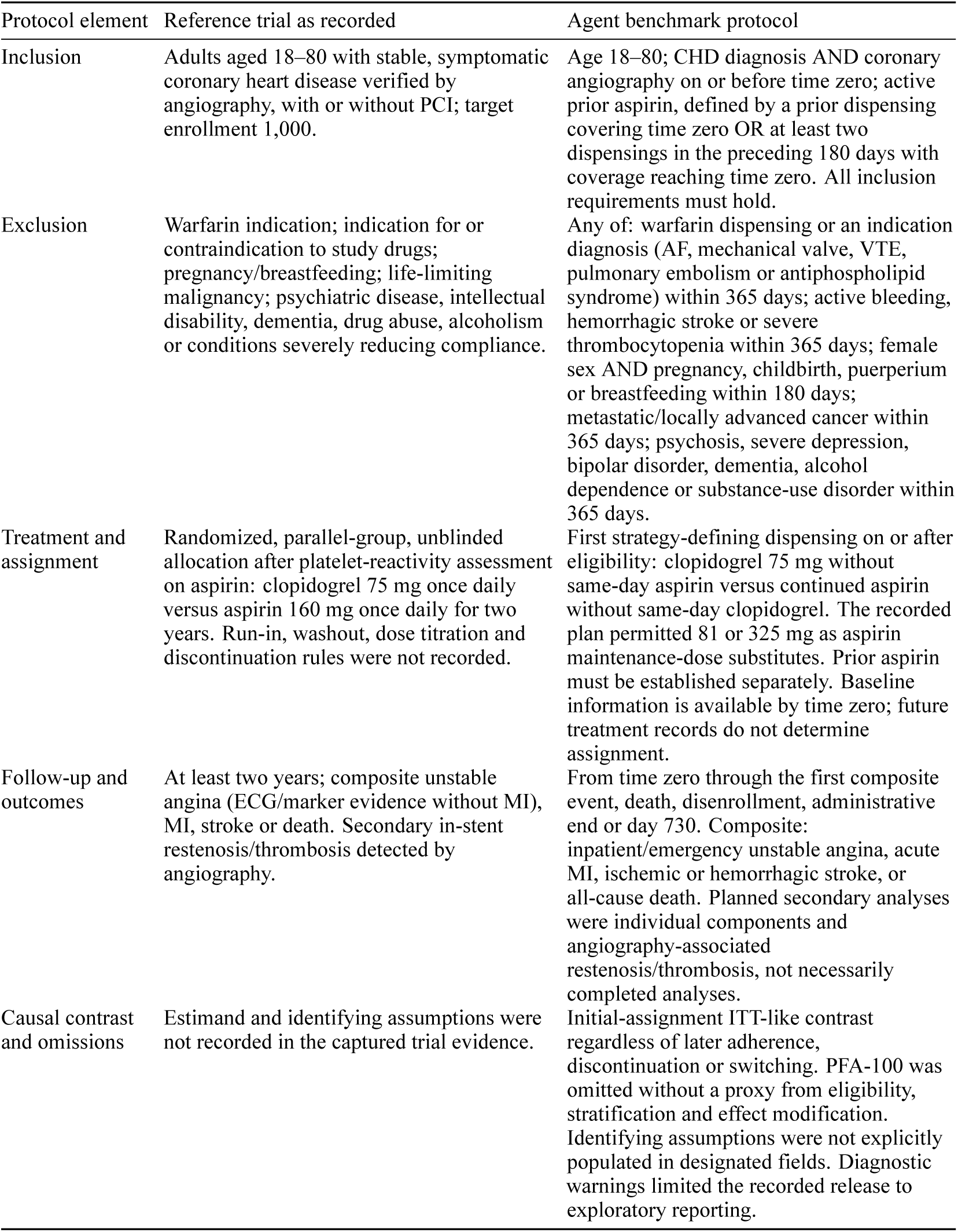
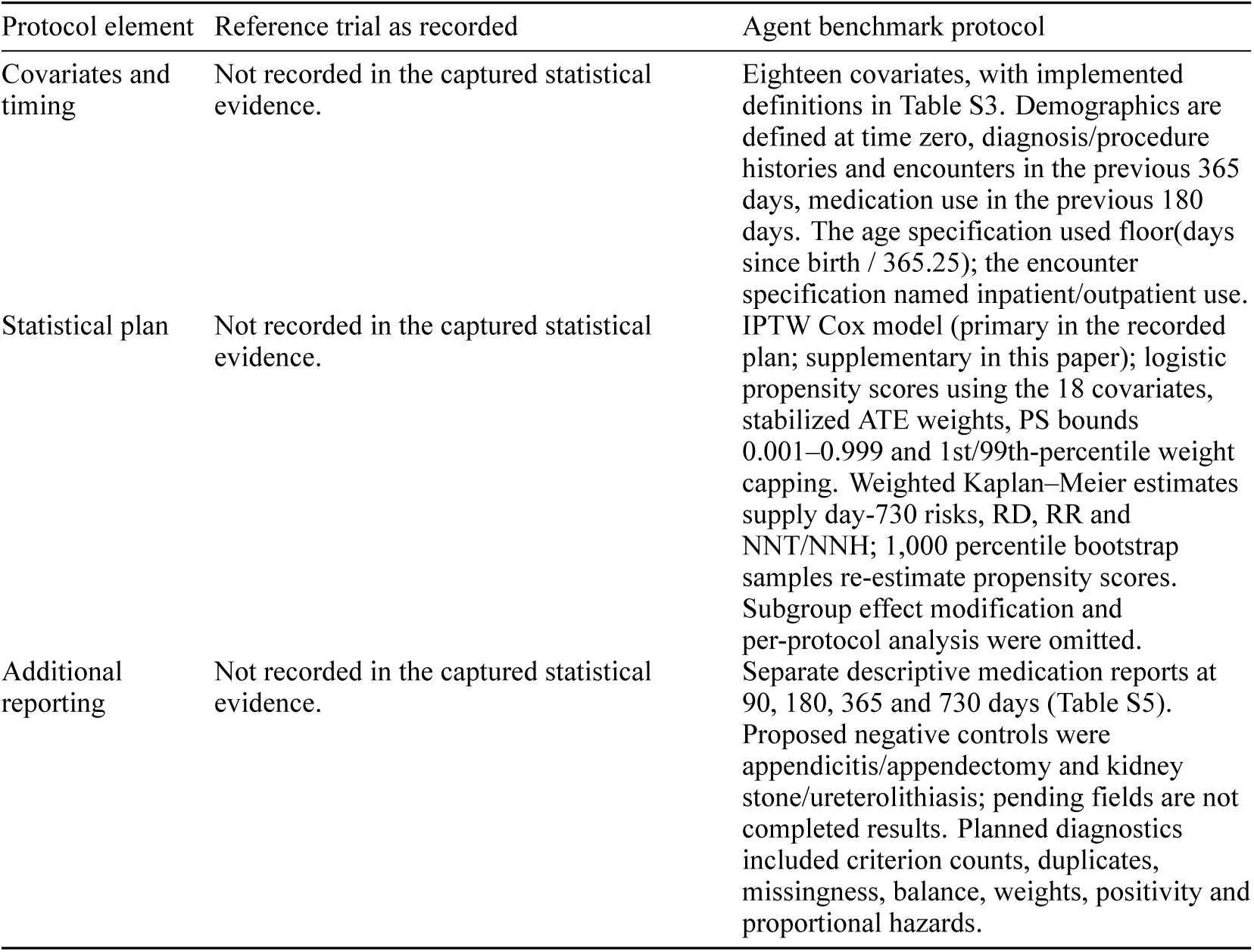

Dose equivalence was not established (main Section 3.1). The broad stroke wording and required angiography differed from parts of the executed definitions. Hyperlipidemia diagnoses and encounters represent recorded lipid-related risk and healthcare use, not measured lipids or adherence.

#### Older-age changes only

The agent changed the age bounds to 81–90 and the requested aspirin label to 162 mg/day, retaining the benchmark definitions and accepted 81-mg product proxy without a tablet-count requirement. The same covariates, windows, outcomes, omission decisions, initial-assignment analysis and medication-reporting rules applied. The CHD wording differed on whether evidence could occur on time zero or had to precede it; the encounter source description also covered all settings rather than only inpatient/outpatient encounters.

### S2. Human emulation specification

The human mapping describes the final implemented analysis, not a prospectively approved protocol. Pre-index windows exclude day 0 and include both stated endpoints. All inclusion conditions and none of the exclusion conditions are required.

| Component | Human benchmark implementation |
| --- | --- |
| Candidates, observation and age | Dispensing decision dates among mapped aspirin users. Age 18–80 in completed years; missing birth date fails and death strictly before index excludes. Same-day death is retained. No enrollment eligibility filter; merge overlapping/adjacent spans and attach the span covering index when available. A 365-day baseline enrollment flag is descriptive only. |
| Stable coronary heart disease | Prior CHD diagnosis AND outpatient CHD record AND coronary procedure evidence, all strictly before index, with no mapped acute coronary syndrome in days –90 to –1. Prior history uses all available records. Outpatient/procedure evidence does not establish symptoms or a positive angiogram; the mapped open coronary dilation does not prove PCI. |
| Prior aspirin and P2Y12 washout | Prior mapped aspirin supply must cover day -1, using either of the two mapped aspirin products at any recorded dose. Supply ends at dispensing plus days supplied minus one; no grace or stockpiling, and missing/nonpositive supply cannot establish active use. No P2Y12 dispensing during days -180 to -1 or older P2Y12 supply overlapping that window. Only clopidogrel was observed in the searched P2Y12 inventory; uncaptured treatment remains unavailable. |
| Warfarin exclusion | Prior warfarin supply covering index OR warfarin dispensing in days -90 to -1 together with an AF/VTE indication in days -365 to -1. Untreated indications may be missed; other anticoagulants do not automatically trigger exclusion. |
| Study-drug suitability exclusion | Bleeding in days -30 to -1; any prior hemorrhagic stroke; thrombocytopenia, liver disease or coronary stenting in days -365 to -1; or acute coronary syndrome in days -90 to -1. Recent stenting proxies a competing dual-antiplatelet indication. Allergy, laboratory severity and platelet values are unavailable. |
| Other exclusions | Pregnancy/lactation was unavailable and assigned flag 0, not confirmed absent. Metastatic-disease codes OR mapped cancer-treatment procedures within 365 days proxy life-limiting malignancy, but treatment alone does not establish prognosis. Dementia requires at least one inpatient or two distinct outpatient dates within 365 days; this does not measure adherence or recover unrecorded psychiatric/social conditions. |
| Assignment and dose | Earliest fully eligible dispensing date: aspirin consistent with 81 mg/day and no same-day P2Y12, or clopidogrel 75 mg/day and no same-day aspirin/other P2Y12. One aspirin product was explicitly assigned one 81-mg tablet daily, overriding the source two-tablet regimen; the other retained source-derived dose. Otherwise daily dose uses strength × tablets/day, or quantity/days supplied. Residual or OTC aspirin may remain after clopidogrel initiation. |
| Follow-up and outcome | Earliest composite event, day 730, end of the index-covering enrollment span if available, or provisional cutoff 2034-03-06. Without a covering span, use the cutoff and flag observation uncertainty. Index after cutoff yields unavailable follow-up, not event-free status. Include index-day and day-730 events within bounds; later switching/discontinuation does not censor. Composite: ED/IP acute MI, ED/IP ischemic stroke, unstable angina requiring principal diagnosis without MI in the same encounter, or death. Hemorrhagic stroke/TIA are not stroke outcomes. ECG/biomarkers and within-day ordering are unavailable. |
| Covariates | Table S3 lists the 18 covariates. Histories/encounters use days -365 to -1; medications use days -180 to -1. Diagnosis dates fall back to admission dates, procedures use recorded dates. No matching record yields zero, not verified absence. Missing demographic categories become Unknown; other numeric covariates must be present, with no silent complete-case exclusion or imputation. |
| Causal contrast and weighting | Initial-assignment marginal ATE, clopidogrel A=1 versus aspirin A=0. Main-effects logistic model, age/encounters linear, clinical flags binary and demographics categorical. Unstabilized ATE weights are $1/PS$ and $1/(1-PS)$, with no clipping, truncation, calendar adjustment/restriction or censoring weighting. Report constant or linearly dependent terms. |
| Estimation and uncertainty | Weighted Kaplan–Meier day-730 risks, clopidogrel-minus-aspirin RD, RR and NNT/NNH. Exactly 1,000 patient bootstraps refit propensity scores/weights/curves (seed 20260910, percentile quantile type 1); all succeeded. Failures would withhold affected intervals, not be replaced. RD intervals crossing zero produce separate benefit/harm ranges to infinity. Secondary weighted Cox uses treatment alone, Efron ties and patient-cluster robust variance, not bootstrap HR uncertainty. |
| Assumptions and scope | Requires measured-confounder adjustment, treatment overlap, suitable independent censoring and day-730 support; no survival extrapolation beyond support. Recorded dose, history, symptoms, angiography, ingestion and aspirin cessation remain measurement limitations. Separate restenosis/thrombosis, adherence-effect and medication-report analyses were not supplied. Platelet testing was not required. |

The human extension changed only eligibility to ages 81–90, refitted the propensity model, and used its own benchmark with the same ASCET reference for calibration. Other definitions were unchanged.

### S3. Diagnostic and result tables

**Table S1.** Weight and propensity-score diagnostics: agent and human benchmark and extension.

| Measure | Agent 18–80 | Human 18–80 | Agent 81–90 | Human 81–90 |
| --- | --- | --- | --- | --- |
| Patients, aspirin / clopidogrel | 3,827 / 2,242 | 3,278 / 2,021 | 1,234 / 368 | 1,007 / 479 |
| Minimum weight | 0.6443 | 1.000000147 /<br>1.000000088 | 0.348531 | 1.000000038 /<br>1.000000023 |
| Weight 1st percentile | 0.6449 | Not reported | 0.348531 | Not reported |
| Median weight | 0.9544 | Not reported | 0.855307 | Not reported |
| Mean weight | 0.9963 | Not reported | 0.962842 | Not reported |
| Weight 99th percentile | 1.5112 | 3.2015 / 4.3515 | 2.454767 | 3.938960 /<br>6.461040 |
| Maximum weight | 1.5112 | 5.2813 / 4.8676 | 2.454816 | 6.115294 /<br>7.449165 |
| Effective sample size | 5,829.2 | 2960.9 / 1839.9 | 1,384.6 | 853.6 / 390.6 |
| ESS / patients, % | 96.05% | 90.33 / 91.04 | Not reported | Not reported |
| Minimum propensity score | 0.0010 / 0.2362 | 1.4672234e-7 /<br>0.20544168 | 0.001000 /<br>0.046181 | 0.00000003768 /<br>0.1342432308 |
| Propensity-score 1st percentile | 0.1210 / 0.2468 | Not reported | 0.010229 /<br>0.060621 | Not reported |
| Median propensity score | 0.3417 / 0.3997 | Not reported | 0.120594 /<br>0.360983 | Not reported |
| Propensity-score 99th percentile | 0.5847 / 0.5933 | Not reported | 0.669643 /<br>0.771752 | Not reported |
| Maximum propensity score | 0.6702 / 0.6683 | 0.810651021 /<br>0.999999912 | 0.784313 /<br>0.786396 | 0.8364755691 /<br>0.9999999771 |
| Patients at PS lower boundary 0.001 | 4 / 0 | Not clipped | 2 / 0 | Not clipped |
| Patients at PS upper boundary 0.999 | 0 / 0 | Not clipped | 0 / 0 | Not clipped |

Paired entries are aspirin / clopidogrel. Agent weight summaries and ESS are pooled; human weight summaries/ESS and all PS entries are arm-specific. Agent weights are stabilized/capped after PS clipping; human weights are unstabilized, uncapped and unclipped. ESS = (sum of weights)² / sum of squared weights, not patients retained.

The agent older-age analysis had zero null patient IDs, zero duplicate patient rows, and no missing values in 24 constructed fields. Human older-age duplicate rows were zero; null-ID and field-level missingness summaries were not separately tabulated. These checks do not establish complete source capture. The human analysis passed 14 structural/temporal checks and 1,000 bootstrap draws; these were not an independent reimplementation of the agent analysis.

**Table S2.**
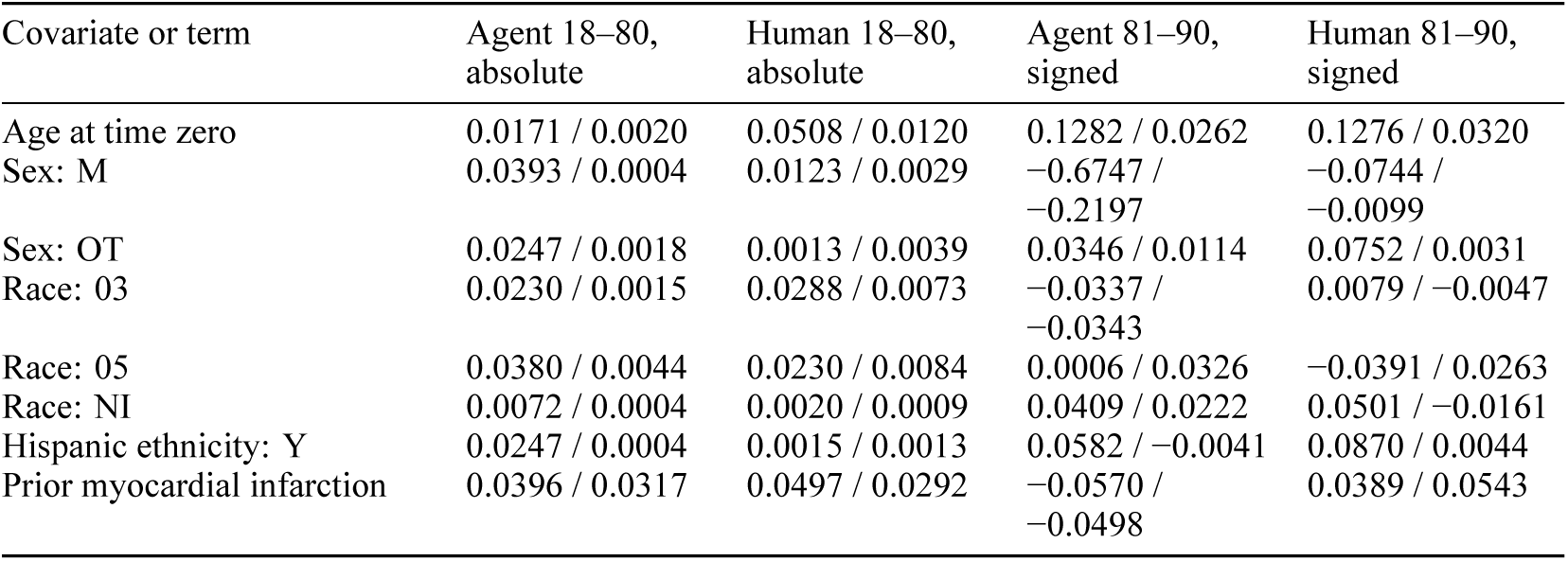
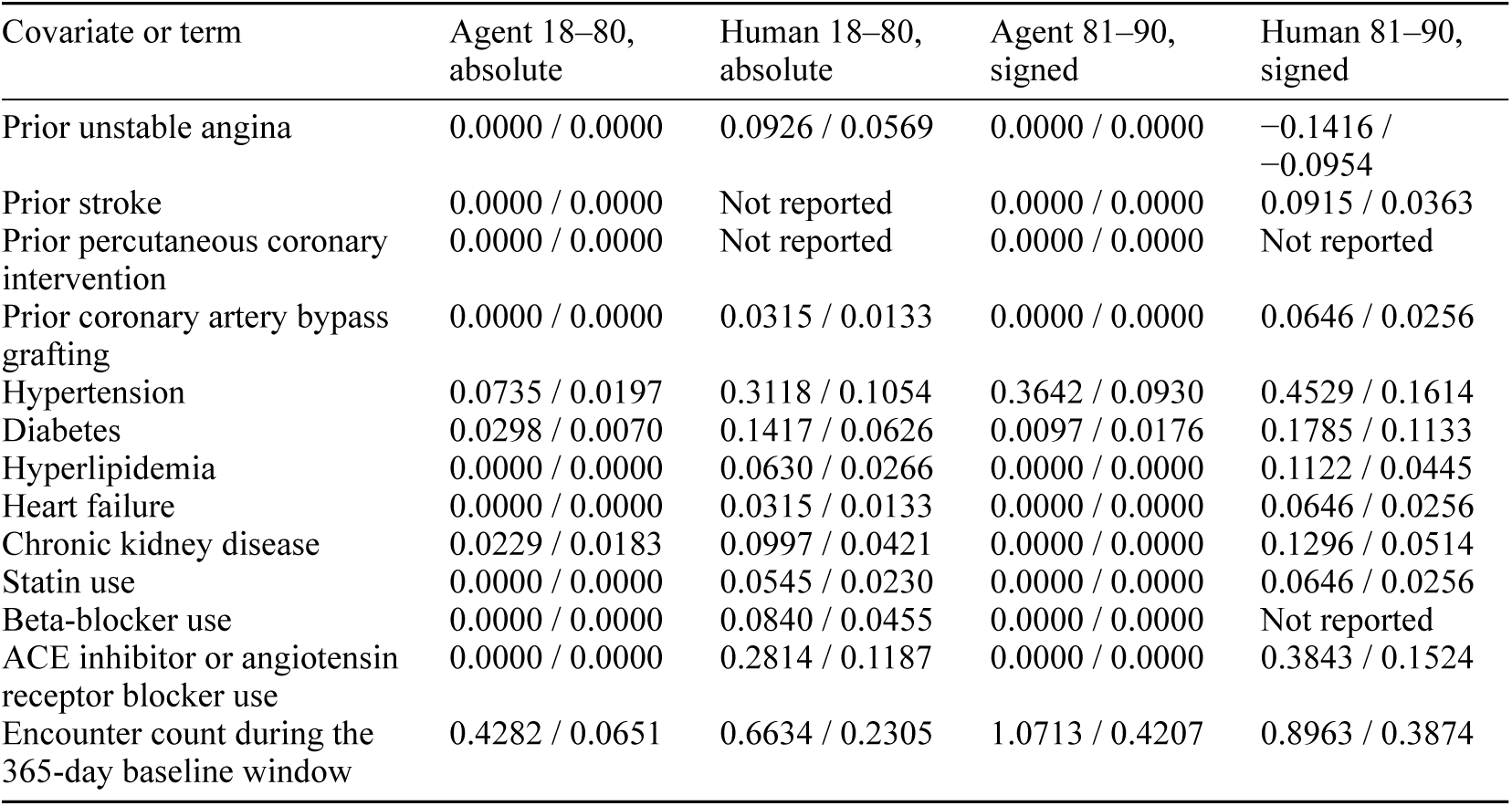
Covariate balance: before / after weighting.

| Covariate or term | Agent 18–80, absolute | Human 18–80, absolute | Agent 81–90, signed | Human 81–90, signed |
| --- | --- | --- | --- | --- |
| Age at time zero | 0.0171 / 0.0020 | 0.0508 / 0.0120 | 0.1282 / 0.0262 | 0.1276 / 0.0320 |
| Sex: M | 0.0393 / 0.0004 | 0.0123 / 0.0029 | −0.6747 /<br>−0.2197 | −0.0744 /<br>−0.0099 |
| Sex: OT | 0.0247 / 0.0018 | 0.0013 / 0.0039 | 0.0346 / 0.0114 | 0.0752 / 0.0031 |
| Race: 03 | 0.0230 / 0.0015 | 0.0288 / 0.0073 | −0.0337 /<br>−0.0343 | 0.0079 / −0.0047 |
| Race: 05 | 0.0380 / 0.0044 | 0.0230 / 0.0084 | 0.0006 / 0.0326 | −0.0391 / 0.0263 |
| Race: NI | 0.0072 / 0.0004 | 0.0020 / 0.0009 | 0.0409 / 0.0222 | 0.0501 / −0.0161 |
| Hispanic ethnicity: Y | 0.0247 / 0.0004 | 0.0015 / 0.0013 | 0.0582 / −0.0041 | 0.0870 / 0.0044 |
| Prior myocardial infarction | 0.0396 / 0.0317 | 0.0497 / 0.0292 | −0.0570 /<br>−0.0498 | 0.0389 / 0.0543 |

| Covariate or term | Agent 18–80,<br>absolute | Human 18–80,<br>absolute | Agent 81–90,<br>signed | Human 81–90,<br>signed |
| --- | --- | --- | --- | --- |
| Prior unstable angina | 0.0000 / 0.0000 | 0.0926 / 0.0569 | 0.0000 / 0.0000 | –0.1416 /<br>–0.0954 |
| Prior stroke | 0.0000 / 0.0000 | Not reported | 0.0000 / 0.0000 | 0.0915 / 0.0363 |
| Prior percutaneous coronary<br>intervention | 0.0000 / 0.0000 | Not reported | 0.0000 / 0.0000 | Not reported |
| Prior coronary artery bypass<br>grafting | 0.0000 / 0.0000 | 0.0315 / 0.0133 | 0.0000 / 0.0000 | 0.0646 / 0.0256 |
| Hypertension | 0.0735 / 0.0197 | 0.3118 / 0.1054 | 0.3642 / 0.0930 | 0.4529 / 0.1614 |
| Diabetes | 0.0298 / 0.0070 | 0.1417 / 0.0626 | 0.0097 / 0.0176 | 0.1785 / 0.1133 |
| Hyperlipidemia | 0.0000 / 0.0000 | 0.0630 / 0.0266 | 0.0000 / 0.0000 | 0.1122 / 0.0445 |
| Heart failure | 0.0000 / 0.0000 | 0.0315 / 0.0133 | 0.0000 / 0.0000 | 0.0646 / 0.0256 |
| Chronic kidney disease | 0.0229 / 0.0183 | 0.0997 / 0.0421 | 0.0000 / 0.0000 | 0.1296 / 0.0514 |
| Statin use | 0.0000 / 0.0000 | 0.0545 / 0.0230 | 0.0000 / 0.0000 | 0.0646 / 0.0256 |
| Beta-blocker use | 0.0000 / 0.0000 | 0.0840 / 0.0455 | 0.0000 / 0.0000 | Not reported |
| ACE inhibitor or angiotensin<br>receptor blocker use | 0.0000 / 0.0000 | 0.2814 / 0.1187 | 0.0000 / 0.0000 | 0.3843 / 0.1524 |
| Encounter count during the<br>365-day baseline window | 0.4282 / 0.0651 | 0.6634 / 0.2305 | 1.0713 / 0.4207 | 0.8963 / 0.3874 |

Each cell is before / after weighting. Benchmark values are absolute SMDs; extension values are signed. The agent’s 18 covariates yielded 21 terms. Nine benchmark covariates were zero for every patient: prior unstable angina, stroke, PCI, CABG, hyperlipidemia, heart failure, statin, beta-blocker and ACE inhibitor/ARB use. The extension also had constant-zero chronic kidney disease. Their reported 0.0000 SMDs are not evidence of successful confounding adjustment.

Human benchmark prior stroke/PCI and extension prior PCI/beta-blocker use had no reported SMD because they were constant. Extension statin use was omitted from the propensity model for linear dependence but remains in descriptive balance. Human SMDs use the fixed unweighted pooled within-arm SD. After weighting, agent extension male sex and encounters, human benchmark hypertension/ACE inhibitor or ARB use/encounters, and human extension hypertension/diabetes/ACE inhibitor or ARB use/encounters exceeded absolute SMD 0.10. The respective extension maxima were 0.421 and 0.387, with 21 and 19 reported terms.

**Table S3.** Implemented baseline covariate definitions: agent and human.

| Covariate | Agent implementation | Human implementation |
| --- | --- | --- |
| Age at time zero | Completed calendar years from birth date to index using birthday comparison. | Completed years from birth date to index |
| Sex | Recorded sex; missing values replaced with the unknown category. | Recorded sex |
| Race | Recorded race, unchanged. | Recorded race |
| Hispanic ethnicity | Recorded Hispanic ethnicity, unchanged. | Recorded Hispanic ethnicity |
| Prior myocardial infarction | Diagnosis flag using the saved prior-MI definition. | ICD-10-CM I21.4, I21.9, I25.2; ICD-9-CM 410.90, 412 |
| Prior unstable angina | Diagnosis flag using the saved prior-unstable-angina definition. This set contains only ICD-9-CM 411.1. | ICD-10-CM I20.0 |
| Prior stroke | Diagnosis flag using the complete saved prior-stroke history set. | ICD-10-CM I63.9, I61.9, I60.9; ICD-9-CM 434.91 |
| Prior PCI | Procedure flag using the five saved prior-PCI procedure selectors. | CPT 92928 |
| Prior CABG | Procedure flag using the nine saved prior-CABG procedure selectors. | CPT 33533 |
| Hypertension | Diagnosis flag using the saved hypertension definition. | ICD-10-CM I10; ICD-9-CM 401.9 |
| Diabetes | Diagnosis flag using the saved diabetes definition. | ICD-10-CM E11.9, E11.319; ICD-9-CM 250.00 |
| Hyperlipidemia | Diagnosis flag using the saved hyperlipidemia definition. | ICD-10-CM E78.5 |
| Heart failure | Diagnosis flag using the saved heart-failure definition. | ICD-10-CM I50.9, I50.22; ICD-9-CM 428.0 |
| Chronic kidney disease | Diagnosis flag using the saved chronic-kidney-disease definition. | ICD-10-CM N18.31, N18.4, N18.5, N18.6; ICD-9-CM 585.9 |
| Statin use | At least one dispensing matching the three saved statin NDCs. | Atorvastatin dispensing, NDC 72162155609 |
| Beta-blocker use | At least one dispensing matching the 65 saved beta-blocker NDCs; saved product descriptions include non-beta-blocker drugs. | Carvedilol dispensing, NDC 00007414120 |
| ACE inhibitor/ARB use | At least one dispensing matching the three saved ACE inhibitor/ARB NDCs; saved product descriptions identify angiotensin II rather than ACE inhibitors/ARBs. | Any mapped lisinopril, ramipril, losartan or telmisartan dispensing |
| Number of encounters | Distinct encounters dated by admission, with discharge-date fallback, across all settings; no match yields zero. | Distinct encounters by admission date |

All clinical history windows are days −365 to −1 and medication flags use days −180 to −1 unless stated. Both workflows use dispensing flags rather than requiring active supply at index. Agent diagnosis dates have no admission-date fallback; code systems are matched separately. No match yields zero, not confirmed absence. Agent age uses completed calendar years rather than the protocol’s day-based formula; encounters include all settings. Some beta-blocker and ACE inhibitor/ARB product descriptions conflict with the intended labels.

**Table S4.** Human calendar entry and follow-up in the benchmark and extension.

Panel A. Calendar year of the earliest eligible index date; entries are distinct patients.
| Entry year | Human 18–80 aspirin | Human 18–80 clopidogrel | Human 81–90 aspirin | Human 81–90 clopidogrel |
| --- | --- | --- | --- | --- |
| 2021 | 0 | 4 | 0 | 3 |
| 2022 | 0 | 12 | 0 | 4 |
| 2023 | 0 | 23 | 0 | 5 |
| 2024 | 0 | 28 | 0 | 19 |
| 2025 | 0 | 10 | 0 | 2 |
| 2029 | 139 | 0 | 133 | 0 |
| 2030 | 2,217 | 1,944 | 575 | 446 |
| 2031 | 892 | 0 | 277 | 0 |
| 2032 | 30 | 0 | 22 | 0 |

Panel B. Primary-outcome follow-up.
| Measure | Human 18–80 aspirin | Human 18–80 clopidogrel | Human 81–90 aspirin | Human 81–90 clopidogrel |
| --- | --- | --- | --- | --- |
| Patients, n | 3,278 | 2,021 | 1,007 | 479 |
| First composite events, n | 345 | 178 | 99 | 54 |
| Event-free censoring before day 730, n (%) | 339 (10.34%) | 34 (1.68%) | 237 (23.54) | 10 (2.09) |
| Follow-up reaches day 730, n (benchmark definition) | 2,595 | 1,809 | Not reported | Not reported |
| Sum of IPTW reaching day 730 (benchmark definition) | 4358.27 | 4327.05 | Not reported | Not reported |
| Patients at risk at day 730, n (extension definition) | Not reported | Not reported | 671 | 415 |
| Weighted number at risk at day 730 (extension definition) | Not reported | Not reported | 1,088.12 | 1,062.58 |
| Mean event-ending follow-up, days | Not reported | Not reported | 317.9 | 374.8 |
| Mean enrollment/data-loss follow-up, days | Not reported | Not reported | 359.6 | 560.9 |

Counts are unweighted. Weighted support totals are not patient counts. Benchmark “reaches day 730” includes events on that day, not only event-free survivors; extension figures are labeled as at-risk counts. Extension counts ending follow-up at day 730 were also 671 aspirin and 415 clopidogrel. Early censoring was attributed to enrollment end. Calendar year was not adjusted and no censoring weights were used. Both human cohorts had clopidogrel entrants during 2021–2025 without same-year aspirin counterparts, including 77 benchmark patients. Synthetic records include future calendar dates. Agent calendar-entry counts were not tabulated in the supplied appendix.

**Table S5.** Agent adherence, persistence and opposite-drug dispensing.

| Agent cohort and arm | Days | Observable patients, n | Mean PDC | PDC $\geq 0.80$ / persistent, n (%) | Opposite-drug dispensing, n (%) | Median days to opposite drug |
| --- | --- | --- | --- | --- | --- | --- |
| 18–80 Aspirin | 90 | 3,814 | 1.000 | 3,814 (100.0%) / 3,814 (100.0%) | 90 (2.4%) | 45 |
| 18–80 Clopidogrel | 90 | 2,231 | 1.000 | 2,231 (100.0%) / 2,231 (100.0%) | 0 (0.0%) | N/A |
| 18–80 Aspirin | 180 | 3,792 | 0.953 | 3,165 (83.5%) / 3,165 (83.5%) | 623 (16.4%) | 90 |
| 18–80 Clopidogrel | 180 | 2,222 | 1.000 | 2,222 (100.0%) / 2,222 (100.0%) | 0 (0.0%) | N/A |
| 18–80 Aspirin | 365 | 3,756 | 0.890 | 3,114 (82.9%) / 3,114 (82.9%) | 619 (16.5%) | 90 |
| 18–80 Clopidogrel | 365 | 2,209 | 1.000 | 2,209 (100.0%) / 2,209 (100.0%) | 0 (0.0%) | N/A |
| 18–80 Aspirin | 730 | 3,555 | 0.845 | 2,839 (79.9%) / 2,780 (78.2%) | 610 (17.2%) | 90 |
| 18–80 Clopidogrel | 730 | 2,180 | 1.000 | 2,180 (100.0%) / 2,180 (100.0%) | 0 (0.0%) | N/A |
| 81–90 Aspirin | 90 | 1,226 | 1.000 | 1,226 (100.0%) / 1,226 (100.0%) | 17 (1.4%) | 45 |
| 81–90 Clopidogrel | 90 | 367 | 1.000 | 367 (100.0%) / 367 (100.0%) | 0 (0.0%) | N/A |
| 81–90 Aspirin | 180 | 1,212 | 0.922 | 858 (70.8%) / 858 (70.8%) | 345 (28.5%) | 90 |
| 81–90 Clopidogrel | 180 | 365 | 1.000 | 365 (100.0%) / 365 (100.0%) | 0 (0.0%) | N/A |
| 81–90 Aspirin | 365 | 1,190 | 0.808 | 831 (69.8%) / 831 (69.8%) | 344 (28.9%) | 90 |
| 81–90 Clopidogrel | 365 | 363 | 1.000 | 363 (100.0%) / 363 (100.0%) | 0 (0.0%) | N/A |
| 81–90 Aspirin | 730 | 1,052 | 0.724 | 688 (65.4%) / 679 (64.5%) | 337 (32.0%) | 90 |
| 81–90 Clopidogrel | 730 | 358 | 1.000 | 358 (100.0%) / 358 (100.0%) | 0 (0.0%) | N/A |

Each denominator includes only patients fully observable for the window, covering index through days 89, 179, 364 or 729. Medication observation ends at the earliest horizon, death, latest source dispensing date or latest enrollment end spanning index; nonfatal outcomes do not end it. Counts are unweighted, not cumulative-incidence estimates.

PDC is distinct supplied days/window days; coverage resets at actual refill dates, overlaps count once and leftover supply is not carried forward. Uncovered grace days remain uncovered for PDC. Persistence uses a 30-day grace period; discontinuation is its complement. All median PDCs were 1.000, no restarts/return dispensings occurred, and median discontinuation time was 165 days wherever discontinuation occurred. All opposite-drug dispensings overlapped assigned-drug supply and were classified as add-on, not confirmed replacement; hence add-on counts equal those shown. Clopidogrel-arm return percentages were not estimable because no opposite-drug dispensing occurred. Dispensing measures availability, not ingestion. These reports did not alter baseline assignment or censor the main analysis; no human medication reports were supplied.

**Table S6.** Router evaluation: all ten questions, answer key and returned recommendation.

| ID | Exact clinical question submitted | Reference route and accepted trial | Returned route and selected trial |
| --- | --- | --- | --- |
| Q01 | Among adults aged 50 years or older with systolic blood pressure of 130–180 mmHg and elevated cardiovascular risk, but without diabetes or previous stroke, compare treatment targeting systolic blood pressure below 120 mmHg versus below 140 mmHg. Evaluate the first occurrence of myocardial infarction, other acute coronary syndrome, stroke, heart failure, or cardiovascular death. Use electronic health records. | Replication<br>SPRINT<br>(NCT01206062) | Replication<br>SPRINT<br>(NCT01206062) |
| Q02 | Among adults with symptomatic chronic heart failure, NYHA class II–IV, left ventricular ejection fraction of 40% or less, and eGFR of at least 30 mL/min/1.73 m<sup>2</sup>, compare adding dapagliflozin 10 mg once daily to recommended heart-failure therapy versus continuing recommended therapy without an SGLT2 inhibitor. Include patients with and without diabetes. Evaluate cardiovascular death or worsening heart failure requiring hospitalization or urgent intravenous treatment. Use electronic health records. | Replication<br>DAPA-HF<br>(NCT03036124) | Replication<br>DAPA-HF<br>(NCT03036124) |
| Q03 | Among adults aged 45 years or older with BMI of at least 27 kg/m<sup>2</sup>, established cardiovascular disease, and no diabetes, compare initiating once-weekly subcutaneous semaglutide, titrated toward 2.4 mg, versus usual care without a GLP-1 receptor agonist. Evaluate cardiovascular death, nonfatal myocardial infarction, or nonfatal stroke. Use electronic health records linked to claims. | Replication<br>SELECT<br>(NCT03574597) | Replication<br>SELECT<br>(NCT03574597) |
| Q04 | Among adults undergoing elective total knee replacement, compare rivaroxaban versus enoxaparin for preventing postoperative venous thromboembolism. Use claims data. | Replication<br>RECORD3<br>(NCT00361894) | Replication<br>RECORD 3<br>(NCT00361894) |
| Q05 | Among adults aged 55 years or older with established atherosclerotic vascular disease or diabetes with end-organ damage, but without heart failure, compare four separate treatment strategies: ramipril 10 mg/day, telmisartan 80 mg/day, losartan 100 mg/day, and valsartan 160 mg/day. Evaluate the five-year composite risk of cardiovascular death, myocardial infarction, stroke, or hospitalization for heart failure. Estimate risk in each group and risk differences and risk ratios relative to ramipril, with 95% confidence intervals. Use electronic health records linked to claims. | BenchExCal<br>ONTARGET<br>(NCT00153101) | BenchExCal<br>ONTARGET<br>(NCT00153101) |
| Q06 | Among adults aged 45 years or older with BMI of at least 27 kg/m<sup>2</sup> and cardiovascular risk factors, but without diabetes or established cardiovascular disease, compare initiating once-weekly subcutaneous semaglutide, titrated toward 2.4 mg, versus usual care without a GLP-1 receptor agonist. Evaluate cardiovascular death, nonfatal myocardial infarction, or nonfatal stroke. Use electronic health records linked to claims. | BenchExCal<br>SELECT<br>(NCT03574597) | BenchExCal<br>SELECT<br>(NCT03574597) |
| Q07 | Among adults receiving maintenance hemodialysis on a Monday–Wednesday–Friday schedule who miss Monday’s session for a nonmedical reason and have no immediate indication for emergency dialysis, compare providing a make-up session on Tuesday versus resuming dialysis on Wednesday. Evaluate hospitalization or emergency-department attendance during the following seven days. Use linked dialysis records and claims. | Direct emulation<br>No accepted<br>randomized-trial<br>benchmark | Direct emulation<br>None selected |
| Q08 | Among clinically stable adults accepted for non-ICU hospital admission who meet inpatient-hallway eligibility criteria during hospital overcrowding, compare transfer within one hour to a staffed inpatient hallway versus continued emergency-department boarding until a standard inpatient bed becomes available. Evaluate seven-day mortality or unplanned ICU transfer. Use hospital electronic records and bed-management data. | Direct emulation<br>No accepted<br>randomized-trial<br>benchmark | Direct emulation<br>None selected |
| Q09 | Among adults admitted to medical-surgical wards, compare hospitals introducing a minimum staffing policy of no more than four patients per licensed nurse during daytime shifts and seven overnight versus hospitals continuing their existing staffing policy. Evaluate 30-day all-cause mortality. Use linked staffing records, hospital records, and mortality data. | Direct emulation<br>No accepted<br>randomized-trial<br>benchmark | Direct emulation<br>None selected |
| Q10 | Among adults aged 45 years or older with BMI $\geq 27$ kg/m<sup>2</sup>, established cardiovascular disease, and no diabetes, compare initiating oral semaglutide, titrated to 14 mg once daily, versus usual care without a GLP-1 receptor agonist. Estimate the three-year composite risk of cardiovascular death, nonfatal myocardial infarction, or nonfatal stroke, including risk difference and risk ratio with 95% confidence intervals. Use EHR linked to claims. | BenchExCal<br>SELECT preferred<br>(NCT03574597);<br>SOUL<br>(NCT03914326)<br>acceptable with a<br>justified population<br>extension | BenchExCal<br>SELECT<br>(NCT03574597) |

The router used Gemini 3.7 Flash with low reasoning and search access, without the answer key. Recommendations were scored before human confirmation. All ten questions form one development set, not a held-out test. Route agreement was 10/10 (four replication, three BenchExCal, three direct emulation); trial agreement was 7/7 applicable registry identifiers. Trial scoring was N/A for Q07–Q09. No downstream emulation was tested.

Q04 used RECORD3 as its knee-replacement benchmark. RECORD4 is another knee-replacement trial; RECORD1 concerns hip replacement. Q10 accepts SELECT, or SOUL with a justified extension beyond its diabetic population. Direct-emulation labels depend on the focused trial search.

**Table S7.** Additional uncalibrated estimates: agent and human benchmark and extension.

| Measure | Agent 18–80 | Human 18–80 | Agent 81–90 | Human 81–90 |
| --- | --- | --- | --- | --- |
| Aspirin risk, % (95% CI) | 15.43<br>(14.27–16.64) | 11.66<br>(10.42–12.81) | 14.33<br>(12.22–16.43) | 11.33<br>(8.99–13.61) |
| Clopidogrel risk, % (95% CI) | 12.78<br>(11.28–14.20) | 8.96 (7.70–10.27) | 12.15<br>(8.24–16.20) | 11.21<br>(8.18–14.38) |
| Hazard ratio (95% CI) | 0.818<br>(0.707–0.947) | 0.753<br>(0.624–0.909) | 0.840<br>(0.573–1.231) | 0.981<br>(0.683–1.411) |
| Number needed to treat (95% confidence set) | 37.8 (22.1–124.9) | 37.0 (22.3–119.6) | 45.9 (Unbounded benefit/harm confidence set; risk-difference interval crosses zero) | 841.0 (NNT [25.78, infinity] or NNH [26.28, infinity]) |

Main Table 1 reports RD and RR estimates. Risks use weighted Kaplan–Meier estimates through day 730, with patient-bootstrap intervals; HR intervals use robust Cox uncertainty. NNT confidence sets invert RD limits and span benefit/harm when zero is included. All four analyses had 1,000 successful bootstrap samples. Benchmark first-event counts were 571 / 281 (agent) and 345 / 178 (human), in aspirin / clopidogrel order. Later medication changes did not redefine baseline arms.

**Table S8.** Calibration inputs and shared-patient covariance sensitivity: agent and human.

| Calibration quantity | Agent | Human |
| --- | --- | --- |
| Benchmark log risk ratio, B | −0.188226 | Not tabulated here |
| Benchmark log-risk-ratio standard error, sB | 0.070669 | 0.091735 |
| Randomized-reference log risk ratio, R | −0.031746 | Not tabulated here |
| Randomized-reference log-risk-ratio standard error, sR | 0.183721 | 0.183721 |
| Uncalibrated extension log risk ratio, E | −0.164874 | Not tabulated here |
| Extension log-risk-ratio standard error, sE | 0.183825 | Not tabulated here |
| Benchmark divergence, D | −0.156480 | −0.231882 |
| Standard error of benchmark divergence under the working approximation | 0.196844 | Not tabulated here |
| Scaling factor, k | 2.601208 | 1.945283 |
| Scaled divergence mean, kD | −0.407037 | −0.451076 |
| Scaled divergence standard deviation | 0.512031 | 0.399463 |
| Calibrated log risk ratio | 0.242163 | 0.440522 |
| Calibrated log-risk-ratio standard error | 0.544029 | Not tabulated here |
| Benchmark log-risk-ratio variance | Not tabulated here | 0.008415230 |
| Randomized-reference log-risk-ratio variance | Not tabulated here | 0.033753248 |
| Uncalibrated extension log-risk-ratio variance | Not tabulated here | 0.031844301 |
| Primary calibrated log-risk-ratio variance | Not tabulated here | 0.191415186 |
| Shared patients between benchmark and extension, n | 102 | 84 |
| Joint-bootstrap covariance of benchmark and extension log risk ratios | Not tabulated here | −0.000602116 |
| Human shared-patient covariance sensitivity: calibrated RR (95% interval) | Not reported | 1.5535 (0.6556–3.6813) |
| Human shared-patient covariance sensitivity: log-RR variance | Not reported | 0.193757757 |
| Human interval-width variance approximation: RR (95% interval); log-RR variance | Not reported | 1.5159 (0.6679–3.4404);<br>0.174865611 |

B, R and E denote benchmark, randomized-reference and extension log RRs. The saved convention uses D = B − R, k = sE/sB, calibrated log RR = E − kD and working variance sE² + k²(sB² + sR²). Estimated scaling is held fixed and cross-cohort covariance omitted in the primary calculation. Shared patients (102 agent; 84 human) are benchmark–extension overlap, not simultaneous membership in both treatment arms. “Not tabulated here” preserves the differing quantities supplied by the two workflows; it does not mean the quantity is zero.

The human covariance sensitivity used 1,000 joint patient-cluster bootstraps, refitting both propensity models with k fixed. The interval-width calculation approximates variances; neither sensitivity replaces the primary calculation. Both workflows had estimate agreement and standardized-difference agreement with ASCET, but not statistical-significance agreement; standardized differences were 0.794946 (agent) and 1.129205 (human). Estimate agreement places the benchmark point estimate within the reference interval; standardized agreement uses approximately absolute(B − R)/√(sB² + sR²) <1.96. The significance rules differed because the agent also permitted both intervals to include 1. These diagnostics do not establish clinical equivalence or transferability of benchmark divergence.

**Table S9.** Cohort construction: agent and human benchmark and extension.

Panel A. Benchmark ages 18–80.
| Stage (agent / human table IDs) | Agent: total rows (distinct patients) | Agent: rows meeting the criterion (distinct patients) | Human: total rows (distinct patients) | Human: rows meeting the criterion (distinct patients) |
| --- | --- | --- | --- | --- |
| Candidate treatment dates and observation records (T01–T02 / T01–T02) | 595,308 (61,532) | N/A | 124,809 (18,000) | N/A |
| Minimum age $\geq 18$ (T03 / N/A) | 595,308 (61,532) | 595,308 (61,532) | N/A | N/A |
| Maximum age $\leq 80$ (T04 / N/A) | 595,308 (61,532) | 570,272 (60,015) | N/A | N/A |
| Age 18–80 and no recorded pre-index death (N/A / T03) | N/A | N/A | 124,809 (18,000) | 88,419 (11,116) |
| Documented coronary heart disease (T05 / T04) | 595,308 (61,532) | 566,778 (60,476) | 124,809 (18,000) | 74,422 (9,108) |
| Prior aspirin use (T06 / T05) | 595,308 (61,532) | 55,299 (8,000) | 124,809 (18,000) | 71,803 (14,582) |
| Separate P2Y12 washout criterion (N/A / T06) | N/A | N/A | 124,809 (18,000) | 73,299 (18,000) |
| Combined inclusion flags (T07 / T07) | 595,308 (61,532) | 33,549 (6,424) | 124,809 (18,000) | 31,383 (6,535) |
| Warfarin-related exclusion (T08 / T08) | 33,549 (6,424) | 8,214 (2,456) | 31,383 (6,535) | 4,826 (1,848) |
| Study-drug contraindication/suitability exclusion (T09 / T09) | 33,549 (6,424) | 399 (130) | 31,383 (6,535) | 5,965 (2,384) |
| Pregnancy/breastfeeding exclusion (T10 / T10) | 33,549 (6,424) | 0 (0) | 31,383 (6,535) | 0 (0) |
| Malignancy exclusion (T11 / T11) | 33,549 (6,424) | 0 (0) | 31,383 (6,535) | 0 (0) |
| Compliance-related exclusion (T12 / T12) | 33,549 (6,424) | 0 (0) | 31,383 (6,535) | 0 (0) |
| Combined exclusion flags (T13 / T13) | 33,549 (6,424) | 8,461 (2,496) | 31,383 (6,535) | 10,542 (3,539) |
| Final cohort, outcomes, covariates and analysis data (T14–T17 / T14–T17) | 6,069 (6,069) | N/A | 5,299 (5,299) | N/A |

Panel B. Extension ages 81–90.
| Stage (agent / human table IDs) | Agent: total rows (distinct patients) | Agent: rows meeting the criterion (distinct patients) | Human: total rows (distinct patients) | Human: rows meeting the criterion (distinct patients) |
| --- | --- | --- | --- | --- |
| Candidate treatment dates and observation records (T01–T02 / T01–T02) | 595,308 (61,532) | N/A | 124,809 (18,000) | N/A |
| Minimum age $\geq 81$ (T03 / N/A) | 595,308 (61,532) | 25,036 (3,572) | N/A | N/A |
| Maximum age $\leq 90$ (T04 / N/A) | 595,308 (61,532) | 595,254 (61,532) | N/A | N/A |
| Age 81–90 and no recorded pre-index death (N/A / T03) | N/A | N/A | 124,809 (18,000) | 30,594 (5,764) |
| Documented coronary heart disease (T05 / T04) | 595,308 (61,532) | 566,778 (60,476) | 124,809 (18,000) | 74,422 (9,108) |
| Prior aspirin use (T06 / T05) | 595,308 (61,532) | 55,299 (8,000) | 124,809 (18,000) | 71,803 (14,582) |
| Separate P2Y12 washout criterion (N/A / T06) | N/A | N/A | 124,809 (18,000) | 73,299 (18,000) |
| Combined inclusion flags (T07 / T07) | 595,308 (61,532) | 9,824 (1,702) | 124,809 (18,000) | 8,773 (1,741) |
| Warfarin-related exclusion (T08 / T08) | 9,824 (1,702) | 1,086 (354) | 8,773 (1,741) | 440 (167) |
| Study-drug contraindication/suitability exclusion (T09 / T09) | 9,824 (1,702) | 101 (30) | 8,773 (1,741) | 1,562 (589) |
| Pregnancy/breastfeeding exclusion (T10 / T10) | 9,824 (1,702) | 0 (0) | 8,773 (1,741) | 0 (0) |
| Malignancy exclusion (T11 / T11) | 9,824 (1,702) | 0 (0) | 8,773 (1,741) | 0 (0) |
| Compliance-related exclusion (T12 / T12) | 9,824 (1,702) | 0 (0) | 8,773 (1,741) | 0 (0) |
| Combined exclusion flags (T13 / T13) | 9,824 (1,702) | 1,179 (376) | 8,773 (1,741) | 1,989 (712) |
| Final cohort, outcomes, covariates and analysis data (T14–T17 / T14–T17) | 1,602 (1,602) | N/A | 1,486 (1,486) | N/A |
| Agent descriptive coverage and behavior summaries | 1,602 (1,602) | N/A | N/A | N/A |
| Agent descriptive opposite-drug/switch-back events | 2,739 (345) | N/A | N/A | N/A |

Each entry reports total rows and distinct patients. Paired stages with identical counts are identified explicitly; T14–T17 are each one row per final patient. Outcome-table rows are not event counts. Coverage and behavior summaries each have 1,602 rows/patients; switching-event rows differ from the complete-window patient counts in Table S5.

Criterion tables are parallel, not sequential attrition: inclusion-positive means a requirement was met; exclusion-positive means an exclusion was triggered. Counts overlap and cannot be added/subtracted to derive the final cohort. Stage IDs do not necessarily represent the same rule across workflows. The human baseline-enrollment flag was descriptive. Its pregnancy/lactation status was unavailable and set to zero; other zero flags mean no qualifying records among evaluated candidates, not proven absence. N/A is not zero. Arm sizes appear in main Table 1.

**Table S10.**
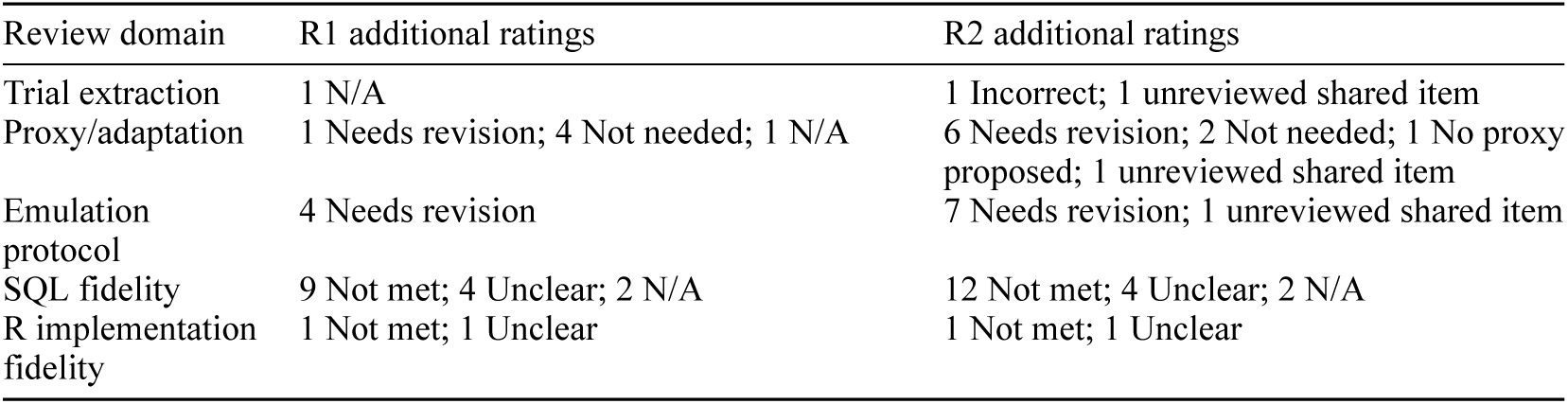
Additional retained reviewer ratings after discussion.

Highest-category scores are in main Table 2. Covariate relevance had no other ratings. SQL/R denominators use Met/(Met + Not met), excluding Unclear and N/A; protocol scores use substantive categories. Overall consistency remained unreviewed and R2’s unmatched prior-clopidogrel item was unscored. Ratings are retained after discussion, not consensus or independent agreement.

